# Prevalence of Risk Factors and Established Cardiovascular Disease Among All of Us Participants: Benchmarking Against National Estimates

**DOI:** 10.1101/2025.07.22.25331985

**Authors:** Mengjing Yang, Tinsae Admassu, Harlan M Krumholz, Chenxi Huang, Karthik Murugiah

## Abstract

**Introduction:** Knowing how All of Us (AoU) Research Program participants differ in demographics and prevalence of cardiovascular risk factors and established cardiovascular disease (CVD) from the U.S population is important for cardiovascular researchers using these data, and to provide context to their findings.

**Methods:** We included AoU participants enrolled between May 2017-June 2022. ‘AoU Survey’ cohort included participants who completed the Personal and Family Health History survey (N=185,232). ‘AoU EHR’ included participants with linked EHR data (N = 287,012). We identified cardiovascular risk factors and established CVD through survey questions in ‘AoU Survey’ and National Health and Nutrition Examination Survey (NHANES), and SNOMED codes for ‘AoU EHR’. Prevalence was compared to the weighted NHANES 2017-March 2020 cycle (N=9,683 representing 248 million), overall and by demographic sub-groups.

**Results:** ‘AoU Survey’ and ‘AoU EHR’ were both older than NHANES (56.1±17 years and 57.6±17 years versus 47.5±18 years) with more female participants. Black participants were underrepresented in ‘AoU Survey’. Prevalence of cardiovascular comorbidities in ‘AoU Survey’ and NHANES were similar overall, but lower when stratified by age and among female participants. Black participants had higher rates of many comorbidities like hypertension, diabetes, coronary artery disease and congestive heart failure than the U.S Black population, but lower chronic kidney disease. ‘AoU EHR’ with its code-based identification had distinctly higher prevalence of most cardiovascular comorbidities.

**Conclusion:** AoU includes more historically underrepresented groups, as intended. However, researchers should understand that demographic composition and disease prevalence change based on data availability and how conditions are ascertained.

## INTRODUCTION

The All of Us (AoU) Research Program, launched by the National Institutes of Health, is a large and diverse U.S. cohort designed to accelerate precision-medicine research.(1) AoU integrates multiple data modalities, including electronic health record (EHR), detailed participant surveys, biospecimens and genomics, physical measurements, and digital health data, while prioritizing enrollment of groups historically underrepresented in biomedical research.(2) For cardiovascular research, these resources can enable new insights into risk factors, disease progression, and treatment responses across diverse communities.

At the same time, AoU enrollment relies on convenience sampling and broad outreach rather than probability-based methods. Consequently, its participants may systematically differ from the general U.S. adult population. Further, AoU analyses may leverage survey responses or linked EHR data, depending on the research question and on data availability. These sources define distinct AoU sub-cohorts whose demographic composition and measured cardiovascular disease (CVD) burden can differ, both from each other and from the U.S. population, potentially affecting the interpretation of study findings. Recognizing these differences can help investigators select the appropriate AoU sub-cohort for a given aim and calibrate expectations about external validity.

Accordingly, this study benchmarks AoU against a national reference by comparing the demographic composition and prevalence of major cardiovascular risk factors and established CVD in AoU sub-cohorts defined by available survey data and by available EHR data with contemporaneous estimates from the National Health and Nutrition Examination Survey (NHANES).(3) We conduct the analyses overall and by key demographic subgroups. Our comprehensive analyses provide critical context for interpreting future cardiovascular research leveraging this landmark cohort.

## METHODS

### Data Sources and Study population

#### AoU Research Program

We used the Controlled Tier Dataset v7 and included participants aged ≥18 years enrolled between May 31, 2017, and June 30, 2022. Cardiovascular comorbidities were identified either from self-reported diagnoses in the Personal and Family Health History (PFHH) survey or from diagnosis codes in linked electronic health records (EHR).

Based on data availability, we defined three analytic cohorts: (1) AoU Overall: all eligible participants regardless of survey/EHR availability; (2) AoU Survey: participants who completed PFHH; and (3) AoU EHR: participants with linked EHR data. Each participant provided written informed consent, and oversight was provided by the AoU Institutional Review Board.

#### NHANES

NHANES is a continuous program conducted by the National Center for Health Statistics using a complex, multistage, stratified, clustered design to produce nationally representative estimates for the U.S. population. All NHANES analyses incorporate provided sample weights and design variables to account for unequal selection probabilities and nonresponse. We analyzed adults (≥18 years) from the 2017–March 2020 cycle who completed the following questionnaires: demographics (DEMO), smoking (SMQ), medical conditions (MCQ), blood pressure (BPQ), kidney conditions (KIQ), occupation (OCQ), health insurance (HIQ), diabetes (DIQ), and health status (HUQ). Use of publicly available, de-identified NHANES data was deemed exempt by the Yale University Institutional Review Board.

### Patient characteristics

#### AoU definitions

Self-reported demographics (e.g., sex, race/ethnicity, employment, income, insurance) were obtained from the “Basics” survey. We examined eight cardiovascular conditions: hypertension (HTN), hyperlipidemia (HLD), diabetes mellitus (DM), coronary artery disease (CAD), acute myocardial infarction (AMI), congestive heart failure (CHF), cerebrovascular accident/transient ischemic attack (CVA/TIA), and chronic kidney disease (CKD). In AoU Survey, a condition was considered ‘Present’ if the participant selected Self to the relevant PFHH item; ‘Absent’ if only family members were selected; ‘Did not answer’ if Skip was selected; and ‘Missing’ if no response was provided. In AoU EHR, a condition was ‘Present’ if at least one corresponding Systematized Nomenclature of Medicine Clinical Terms (SNOMED CT)(4) code appeared any time prior to the participant’s PFHH survey date; otherwise, it was classified ‘Absent’. This approach aligns with the “ever told by a health professional” wording used in survey items and avoids imposing an arbitrary look-back window given heterogeneous EHR observation period. The full SNOMED CT code lists are provided in Supplementary Table 1.

### NHANES definitions

Sociodemographic variables were collected via structured interviews. Cardiovascular comorbidities were defined by self-report using targeted questions: CHF (MCQ.160b), coronary heart disease (MCQ.160c), angina (MCQ.160d), AMI (MCQ.160e), and stroke (MCQ.160f); CKD and dialysis were assessed with KIQ.022 and KIQ.025, respectively; diabetes, hypertension, and hyperlipidemia were identified with DIQ.010, BPQ.020, and BPQ.080. Responses were categorized as ‘Present’ for ‘Yes’, ‘Absent’ for ‘No’, ‘Did not answer’ for ‘Refused’/‘Don’t know’, and ‘Missing’ when no response was recorded.

### Race/ethnicity harmonization

Race and ethnicity were self-reported in both datasets. For cross-dataset comparability, we harmonized categories to Non-Hispanic White, Non-Hispanic Black, Hispanic, Asian, and Other. In NHANES, “Other race including multiracial” aggregates several less common groups without further breakdown. AoU allows more granular self-identification (e.g., American Indian or Alaska Native, Native Hawaiian or other Pacific Islander, Middle Eastern or North African, multiracial). To maintain consistency, these detailed AoU categories were grouped under Other; because the underlying composition differs between datasets, comparisons involving “Other” are not interpreted.

### Statistical analysis

Analyses were conducted in R 4.4.0. For NHANES, we used the survey package to account for the complex design, incorporating weights, strata, and primary sampling units to derive nationally representative estimates and variance. For AoU, which is non-probabilistic, we reported unweighted means, proportions, and 95% confidence intervals.

We first compared key sociodemographic characteristics (age, sex, race/ethnicity, income, insurance) across NHANES and the three AoU cohorts (AoU Overall, AoU Survey, AoU EHR) using two-sample t-tests for continuous variables and chi-square tests for categorical variables. For comorbidity prevalence, comparisons were restricted to NHANES vs AoU Survey and NHANES vs AoU EHR. We further examined group differences within strata defined by age (18-40, 40-65, >65 years), sex, and race/ethnicity using the same framework. Two-sided P<0.05 was considered statistically significant; given the large sample size and multiple comparisons, significance testing was used to highlight patterns rather than to draw definitive causal inferences.

## RESULTS

The AoU Overall cohort included 413,457 participants (Figure 1). Of these, 185,232 (44.8%) completed the PFHH survey (AoU Survey), and 287,012 (69.4%) had linked electronic health records (AoU EHR). The NHANES cohort included 9,683 adults, representing an estimated 248 million U.S. adults after weighting.

**Figure 1:**
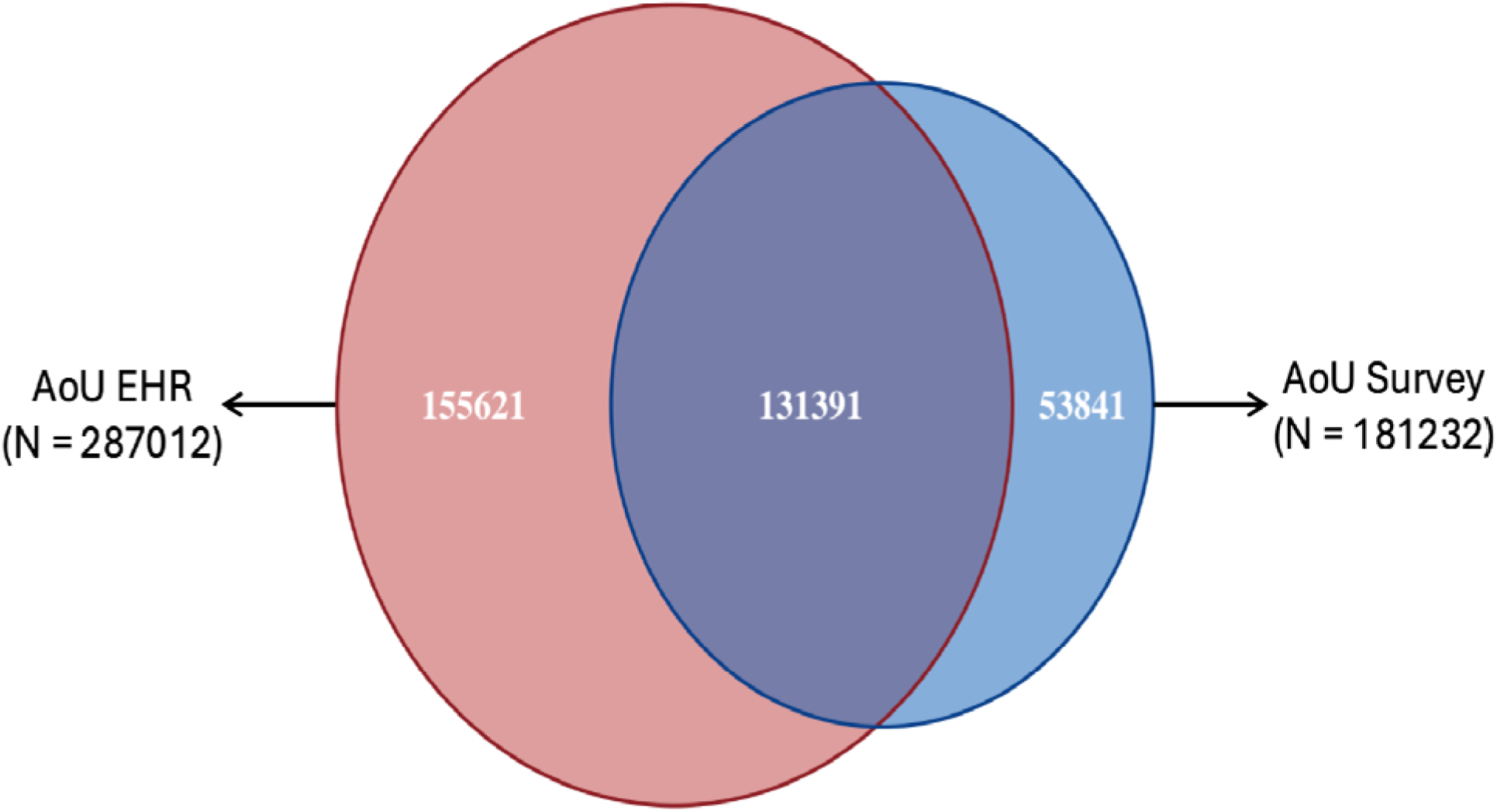
Venn diagram representing the overlap of participants with Person and Family Survey data and EHR data.

Compared with NHANES, AoU participants were older (mean±SD 56.1±17.1 vs 47.5±17.9 years) and more often female (60.4% vs 51.8%), with a higher proportion of Black participants (18.6% vs 11.5%; all P<0.001; Table 1). Both AoU sub-cohorts were also older (AoU Survey 58.1±17.1; AoU EHR 57.6±17.0 years) and had higher percentage of females (AoU Survey 64.0%; AoU EHR 60.1%) than NHANES. Black representation differed by sub-cohort with AoU Survey having 9.1%, lower than NHANES (11.5%) and AoU EHR having 19.9%, higher than NHANES. The higher mean age of AoU Survey versus NHANES was observed across demographic subgroups, with men 62.6 vs 46.7 years in NHANES; women 56.9 vs 48.3; White 61.4 vs 49.8; Black 56.8 vs 45.3; all P<0.001 (Supplementary Table 2).

**Table 1.** Comparison of Socio-Demographic Factors between NHANES and AoU cohorts.

| Socio-demographic Status | NHANES Weighted |  | AoU General<br>(N = 413,457) |  | AoU Survey<br>(N = 185,232) |  | AoU EHR<br>(N = 287,012) |  |
| --- | --- | --- | --- | --- | --- | --- | --- | --- |
| Age (mean, SD, in years) | 47.49 (17.87) |  | 56.06(17.11) |  | 58.14(17.14) |  | 57.63(16.96) |  |
|  | N | % | N | % | N | % | N | % |
| **Age Group** |  |  |  |  |  |  |  |  |
| 18-40 | 94125616 | 37.96 | 91731 | 22.19 | 37045 | 20.00 | 57725 | 20.11 |
| 40-65 | 103189321 | 41.62 | 174886 | 42.30 | 71991 | 38.87 | 120291 | 41.91 |
| >=65 | 50632123 | 20.42 | 146840 | 35.52 | 76196 | 41.14 | 108996 | 37.98 |
| **Sex** |  |  |  |  |  |  |  |  |
| Male | 119592918 | 48.23 | 155169 | 37.53 | 62560 | 33.77 | 108739 | 37.89 |
| Female | 128354142 | 51.77 | 249565 | 60.36 | 118491 | 63.97 | 172401 | 60.07 |
| Did not answer | 0 | 0.00 | 4439 | 1.07 | 1237 | 0.67 | 3169 | 1.10 |
| Missing | 0 | 0.00 | 4284 | 1.04 | 2944 | 1.59 | 2703 | 0.67 |
| **Race/ Ethnicity** |  |  |  |  |  |  |  |  |
| White | 154162965 | 62.18 | 222646 | 53.85 | 129090 | 69.69 | 150402 | 52.40 |
| Black | 28420787 | 11.46 | 77069 | 18.64 | 16918 | 9.13 | 57177 | 19.92 |
| Asian | 14781102 | 5.96 | 13838 | 3.35 | 6086 | 3.29 | 8038 | 2.80 |
| Hispanic/Latino/a/x | 40485465 | 16.33 | 64672 | 15.64 | 17236 | 9.31 | 47699 | 16.62 |
| Other/ >1 | 10096742 | 4.07 | 23443 | 5.67 | 10266 | 5.54 | 15690 | 5.47 |
| Did not answer | 0 | 0.00 | 11692 | 2.83 | 5636 | 3.04 | 8005 | 2.79 |
| Missing | 0 | 0.00 | 97 | 0.02 | 0 | 0.00 | 1 | 0.00 |
| **Marital Status** |  |  |  |  |  |  |  |  |
| Married | 148072184 | 59.72 | 204011 | 49.34 | 107790 | 58.19 | 139534 | 48.62 |
| Widowed, Separated<br>or divorced | 45281859 | 18.26 | 92150 | 22.29 | 35758 | 19.30 | 67086 | 23.37 |
| Never married | 47000622 | 18.96 | 102480 | 24.79 | 36484 | 19.70 | 70297 | 24.49 |
| Did not answer | 94828 | 0.04 | 14719 | 3.56 | 5200 | 2.81 | 10094 | 3.52 |
| Missing | 7497567 | 3.02 | 97 | 0.02 | 0 | 0.00 | 1 | 0.00 |
| **Education (years)** |  |  |  |  |  |  |  |  |
| <12 y | 26538169 | 10.70 | 36432 | 8.81 | 5976 | 3.23 | 27867 | 9.71 |
| 12 y | 64776211 | 26.13 | 77014 | 18.63 | 19654 | 10.61 | 56778 | 19.78 |
| >12 y | 148884269 | 60.05 | 286481 | 69.29 | 154659 | 83.49 | 193184 | 67.31 |
| Did not answer | 2508442 | 0.10 | 13433 | 3.25 | 4943 | 2.67 | 9182 | 3.20 |
| Missing | 7497567 | 3.02 | 97 | 0.00 | 0 | 0.00 | 1 | 0.00 |
| **Self-rated Health** |  |  |  |  |  |  |  |  |
| Good | 201938577 | 81.44 | 293368 | 70.95 | 144907 | 78.23 | 205987 | 71.77 |
| Fair | 39302718 | 15.85 | 81000 | 19.59 | 31524 | 17.02 | 60044 | 20.92 |
| Poor | 6419188 | 2.59 | 20555 | 4.97 | 7352 | 3.97 | 15358 | 5.35 |
| Did not answer | 286576 | 0.12 | 17288 | 4.18 | 1449 | 0.78 | 5467 | 1.90 |
| Missing | 0 | 0.00 | 1246 | 0.30 | 0 | 0.00 | 156 | 0.05 |
| **Health Insurance** |  |  |  |  |  |  |  |  |
| Covered | 214599200 | 86.55 | 371633 | 89.88 | 173855 | 93.86 | 260359 | 90.71 |
| Not covered | 32892008 | 13.27 | 26779 | 6.48 | 6058 | 3.27 | 16733 | 5.83 |
| Did not answer | 455852 | 0.18 | 13158 | 3.18 | 3690 | 1.99 | 8685 | 3.03 |
| Missing | 0 | 0.00 | 1887 | 0.46 | 1629 | 0.88 | 1235 | 0.43 |
| **Smoking** |  |  |  |  |  |  |  |  |
| Yes | 102779026 | 41.45 | 155901 | 37.71 | 65107 | 22.68 | 114595 | 39.93 |
| No | 145104923 | 58.52 | 232630 | 56.26 | 116753 | 40.68 | 164021 | 57.15 |
| Did not answer | 63111 | 0.03 | 23680 | 5.73 | 3372 | 1.17 | 8240 | 2.87 |
| Missing | 0 | 0.00 | 1246 | 0.3 | 0 | 0.00 | 156 | 0.05 |
| **Employment Status** |  |  |  |  |  |  |  |  |
| Employed | 158614770 | 63.97 | 199392 | 48.23 | 98660 | 53.26 | 131320 | 45.75 |
| Unemployed | 89276381 | 36.01 | 197741 | 47.83 | 81166 | 43.82 | 144650 | 50.40 |
| Did not answer | 55909 | 0.02 | 16227 | 3.92 | 5406 | 2.92 | 11041 | 3.85 |
| Missing | 0 | 0.00 | 97 | 0.02 | 0 | 0.00 | 1 | 0.00 |

Relative to AoU EHR, AoU Survey participants were slightly older and more often female and had higher proportions of White participants (69.7% vs 52.4%), greater educational attainment (>12 years: 83.5% vs 67.3%), and higher employment (53.3% vs 45.8%).

The overall prevalence of CV risk factors and established CVD is shown in Figure 2 and Supplementary Table 3. Overall, self-reported prevalence in the AoU Survey cohort were similar to or marginally lower than NHANES estimates (e.g. hypertension 29.6% in AoU Survey and 31.7% in NHANES, DM 11.3% in both, CAD 4.1% in both). In contrast, the AoU EHR cohort with its code-based disease identification had a significantly higher prevalence of all comorbidities except for CVA/TIA, which was lower than in NHANES (e.g. hypertension 41.6%, DM 19.6%, CAD 11.7% etc).

**Figure 2:**
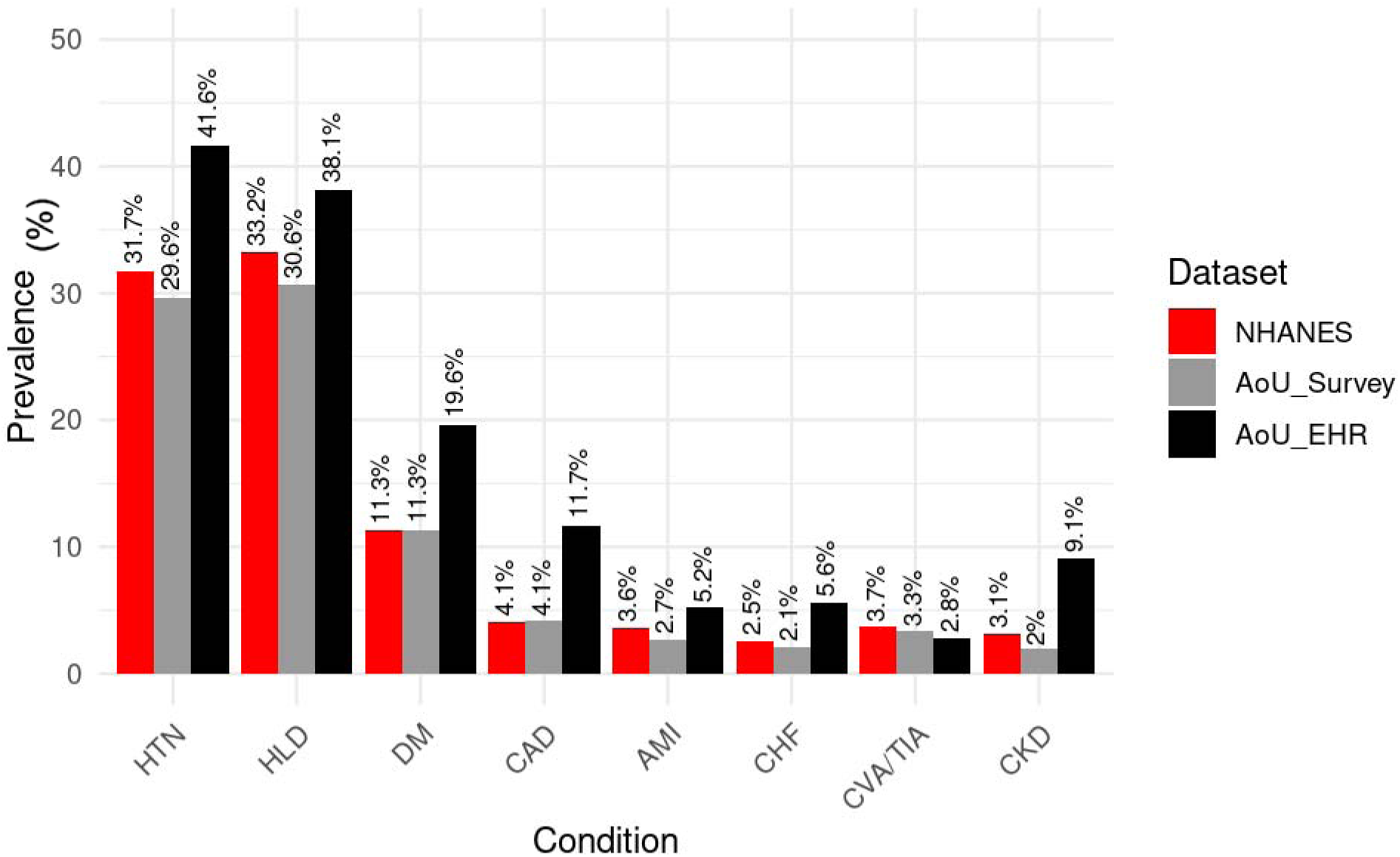
Prevalence of CV risk factors and established CVD. This figure compares the estimated prevalence of major cardiometabolic conditions including acute myocardial infarction (AMI), coronary artery disease (CAD), congestive heart failure (CHF), chronic kidney disease (CKD), cerebrovascular accident/transient ischemic attack (CVA/TIA), diabetes mellitus (DM), hyperlipidemia (HLD), and hypertension (HTN) across three data sources: NHANES (self-report and exam-based), All of Us (AoU) EHR, and AoU survey responses. EHR-based prevalence estimates were consistently higher than survey-based or NHANES estimates for most conditions, particularly for AMI, CAD, and CKD.

The prevalence of CV risk factors and established CVD stratified by age, sex and race/ethnicity is shown in Figures 3 and 4 and Supplementary Table 4. Across age groups, the prevalence of CV risk factors and established CVD was lower in the AoU Survey cohort compared to NHANES, and this pattern was consistent across most comorbidities. A similar pattern was also observed among women. Stratified by race, the prevalence of CV risk factors and established CVD was consistently lower among White participants in the AoU Survey cohort when compared to the White population in NHANES, despite a higher mean age of White participants in AoU. However, Black participants deviated from this pattern, with higher rates of hypertension, diabetes, CAD, CHF, but lower prevalence of CKD.

**Figure 3:**
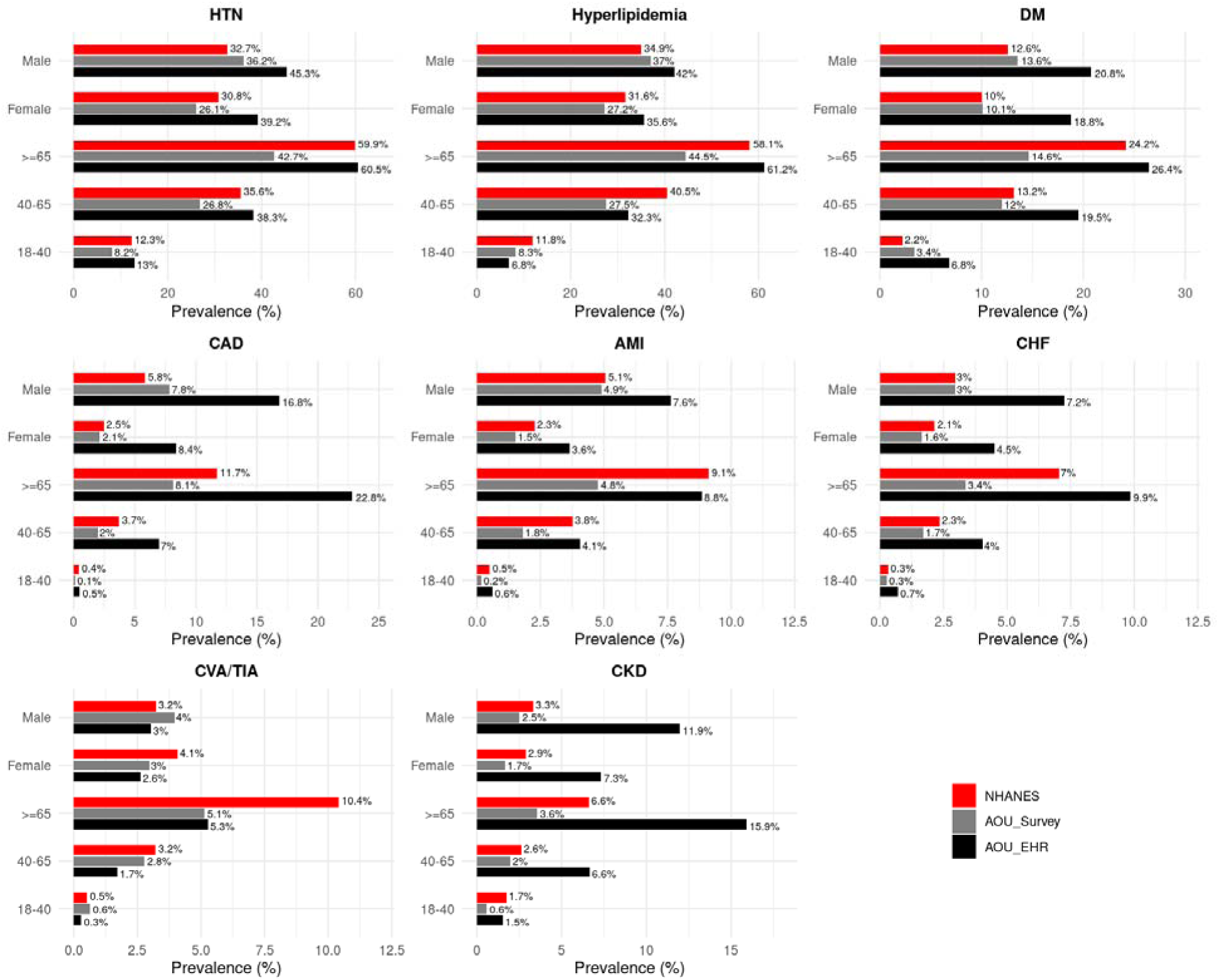
Prevalence of CV risk factors and established CVD by age and sex. This figure presents the prevalence of eight cardiometabolic conditions by demographic subgroups (age groups: 18–40, 40–65, ≥65; and sex: Male, Female) across three datasets. EHR-based data from All of Us generally show higher prevalence rates than self-reported survey responses and NHANES, with the largest age-associated increases observed for hypertension, diabetes, and CAD. The sex-specific trends vary by condition and data source.

**Figure 4:**
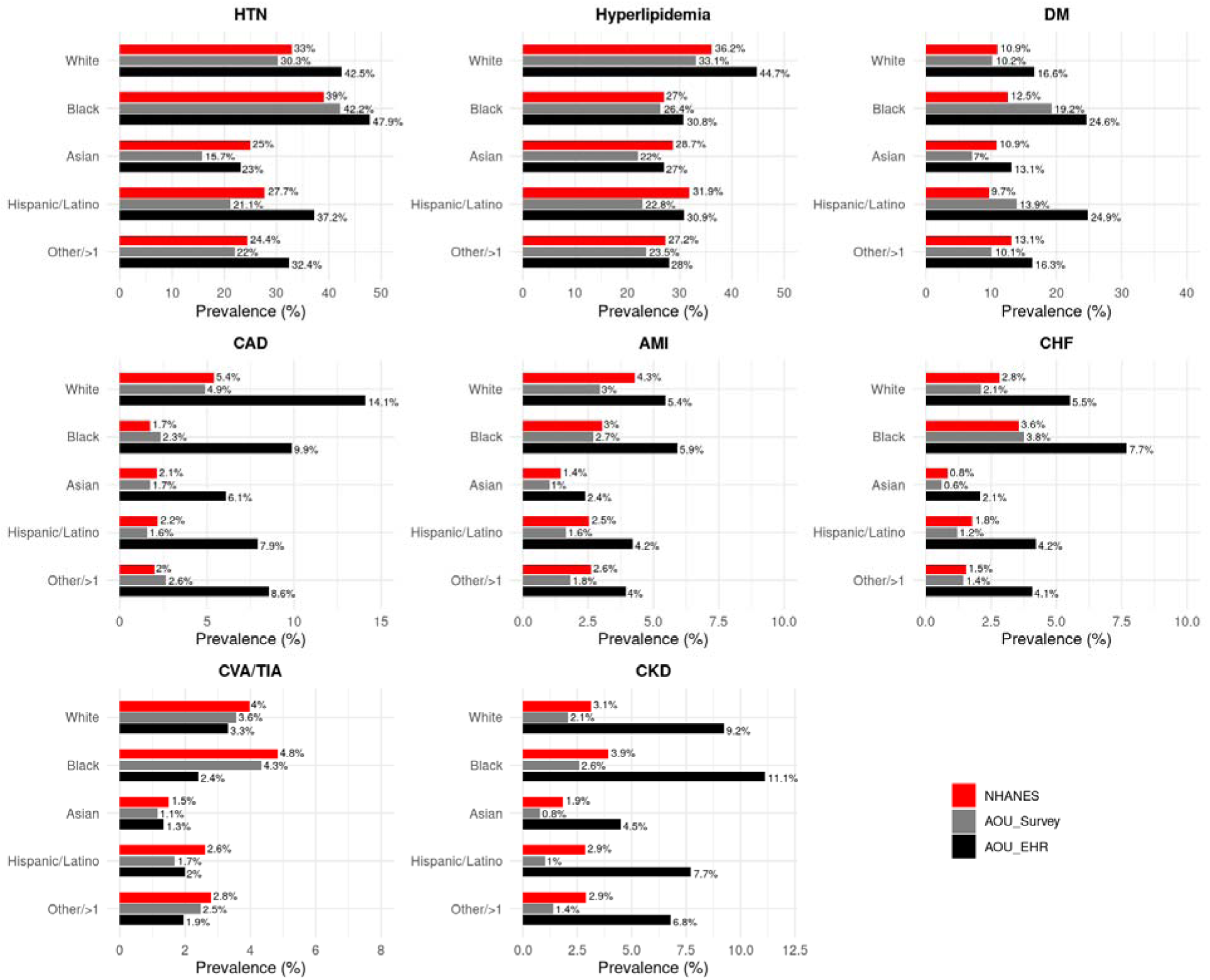
Prevalence of CV risk factors and established CVD by race/ethnicity. This figure shows race/ethnicity-stratified prevalence for key cardiovascular and metabolic conditions, comparing NHANES and both EHR and survey data from All of Us. Disparities in disease burden are evident across racial/ethnic groups, with Black participants generally exhibiting higher prevalence of hypertension and diabetes across all datasets, while Asian participants show lower rates for most conditions.

## DISCUSSION

In this benchmarking study, All of Us (AoU) participants differed from the U.S. adult population in composition, with older age, a higher proportion of women and Black adults, and these differences varied based on participant availability of health survey data and EHR data. CVD burden also differed by ascertainment methods.

Survey-based AoU estimates were broadly similar to national figures overall, but within age strata was consistently lower across most conditions. In contrast, EHR-based prevalence in AoU exceeded both AoU survey and NHANES estimates for most conditions. Subgroup patterns were heterogeneous: women in AoU Survey reported fewer conditions than women in NHANES, whereas Black participants in AoU Survey had higher hypertension, diabetes, and heart failure than Black adults in NHANES.

These findings should be interpreted considering how AoU is assembled and how conditions are captured. AoU is a convenience sample with intentional outreach to groups historically underrepresented in research, which is reflected in a higher overall proportion of Black participants than NHANES; however, representation differs across sub-cohorts, with the AoU Survey cohort in fact including a lower proportion of Black participants than both AoU Overall and NHANES. Differences between the survey-defined and EHR-defined cohorts extended beyond race/ethnicity to education and employment, indicating that choosing a data source effectively selects into different AoU populations. These differences also underscore the persistent socioeconomic and logistic barriers to completing surveys which continue to limit our ability to obtain self-reported health behaviors and health experiences of underrepresented populations.(5) At the same time, disease ascertainment by self-report and clinical coding have their different sets of biases.(6) Self-report can under-ascertain conditions because of recall or health-literacy barriers(7), whereas EHR capture depends on care access, encounter frequency, and coding practices.(8) Our EHR definitions used any prior SNOMED CT code recorded before the participant’s baseline survey date to align conceptually with the “ever diagnosed” wording in both AoU and NHANES surveys, avoiding arbitrary look-back windows in the face of heterogeneous EHR observation periods. Together, these compositional and measurement factors could explain the lower survey-based prevalence within age strata and the higher EHR-based prevalence.

This study adds clarity to the literature(9) by providing a methods-aligned benchmark for AoU CVD burden against a contemporaneous, probability-sampled national reference and by presenting AoU survey-based and EHR-based estimates separately rather than combining sources. Prior AoU comparisons that contrasted EHR-only AoU definitions with NHANES survey estimates(10), or that used non-survey references such as Global Burden of Diseases (GBD)(11), are difficult to interpret because of non-parallel case definitions, broader multimorbidity scopes, or misaligned time windows. By aligning “ever diagnosed” definitions, matching time frames, and stratifying by age, sex, and race/ethnicity, our analysis isolates where AoU diverges most from the national benchmark and shows how those divergences depend on the data source a study elects to use. Relative to legacy cardiovascular cohorts such as MESA(12) and ARIC(13), AoU offers unprecedented scale and multimodal depth; our results clarify how its convenience sampling and multi-source ascertainment should be considered when generalizing AoU-based findings to the U.S. population.

These benchmarks have practical implications. Investigators using AoU for cardiovascular research should prespecify whether analyses will rely on survey or EHR data and recognize that this choice entails different demographic compositions and measured burdens of CVD. Because AoU estimates are unweighted and derived from a convenience sample, they should not be interpreted as national prevalence; instead, contrasts with NHANES can guide expectations about direction and magnitude of differences when translating AoU findings. Stratification by age, sex, and race/ethnicity is essential for interpretation and for assessing whether observed associations are likely to hold in the broader U.S. population.

This work has limitations. It is cross-sectional, using a single survey timepoint and an EHR snapshot, so we do not assess disease onset or progression. Misclassification remains possible in both sources: survey measures may be affected by recall or comprehension, and EHR measures by incomplete capture or coding variability. AoU’s non-probability sampling and unweighted analysis limit direct comparability to NHANES despite the benchmarking approach, and the harmonized “Other” race/ethnicity category differs in composition across datasets and is not interpreted. EHR data are available only for participants who consented and who could be linked through participating healthcare provider organizations; expanding linkage may change the composition of the EHR-defined cohort in future AoU releases. Further, AoU Survey in our study refers to completion of the Personal/Family Health History which was used to identify self-reported comorbidities. Response rates and demographic distributions will vary for other surveys. Finally, we did not examine within-person concordance between survey and EHR because our objective was to compare population burden across data-defined AoU cohorts, not to reconcile sources.

## CONCLUSION

AoU’s breadth and diversity make it a powerful platform for cardiovascular research, but who is included and how conditions are ascertained shape measured CVD burden. By benchmarking AoU survey-defined and EHR-defined cohorts against NHANES using aligned definitions and time frames, we provide actionable context to help researchers choose the appropriate AoU data source for their aims and to judge the generalizability of AoU-based cardiovascular findings.

## Data Availability

All data produced are available online at https://www.researchallofus.org/

## ACKNOWLEDGEMENTS

We gratefully acknowledge All of Us participants for their contributions, without whom this research would not have been possible. We also thank the National Institutes of Health’s All of Us Research Program for making available the participant data examined in this study.

---

**Supplementary Table 1:**
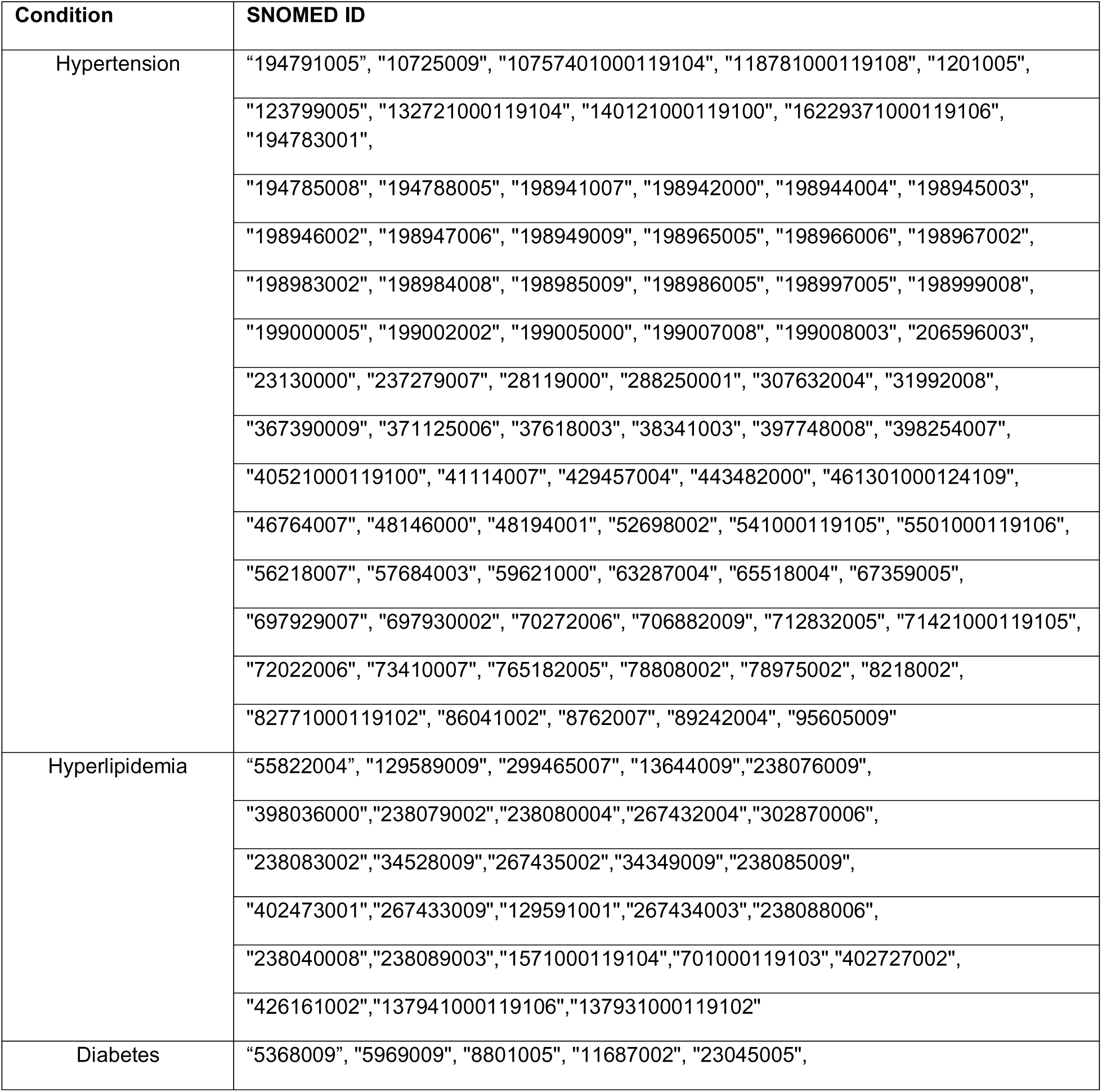
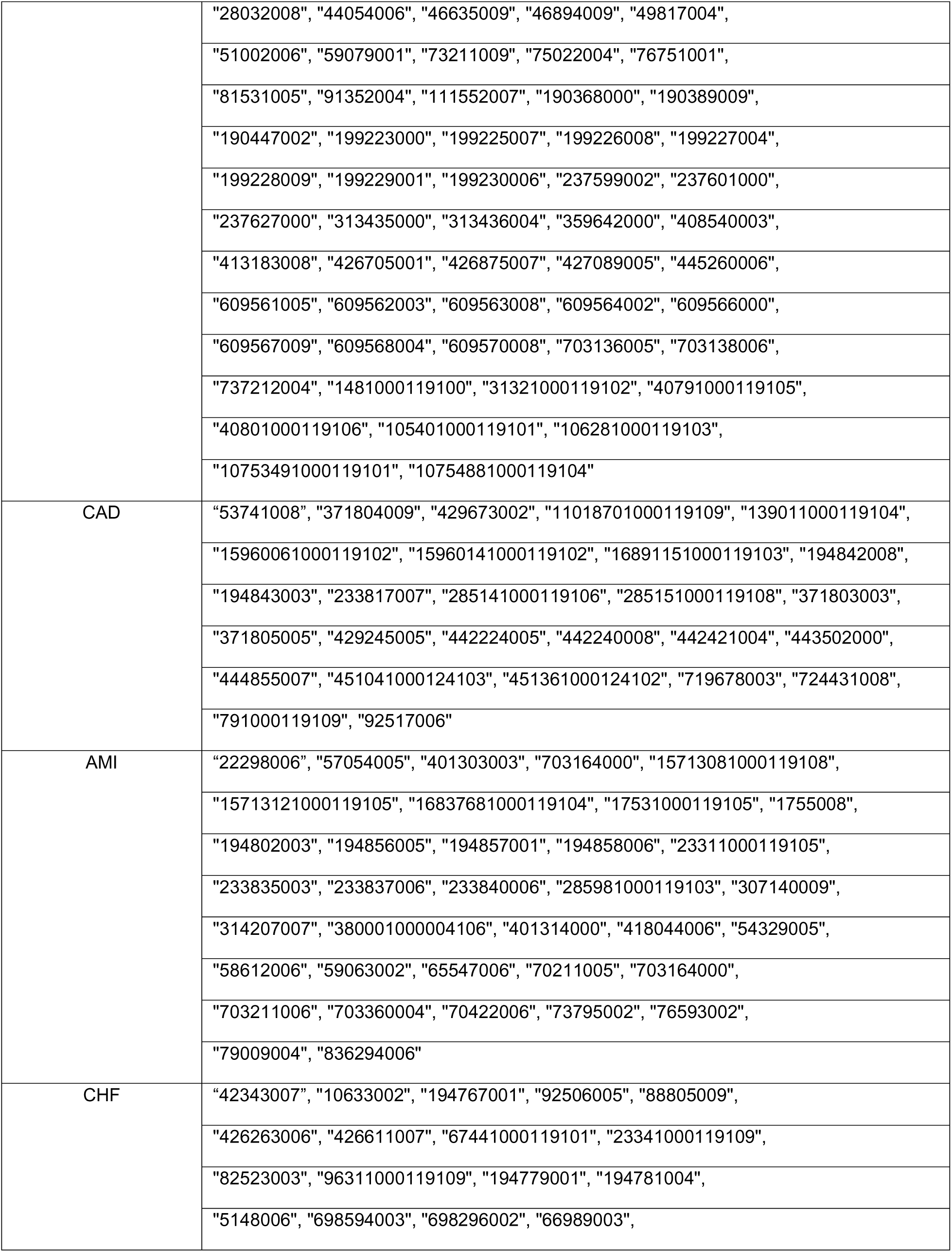
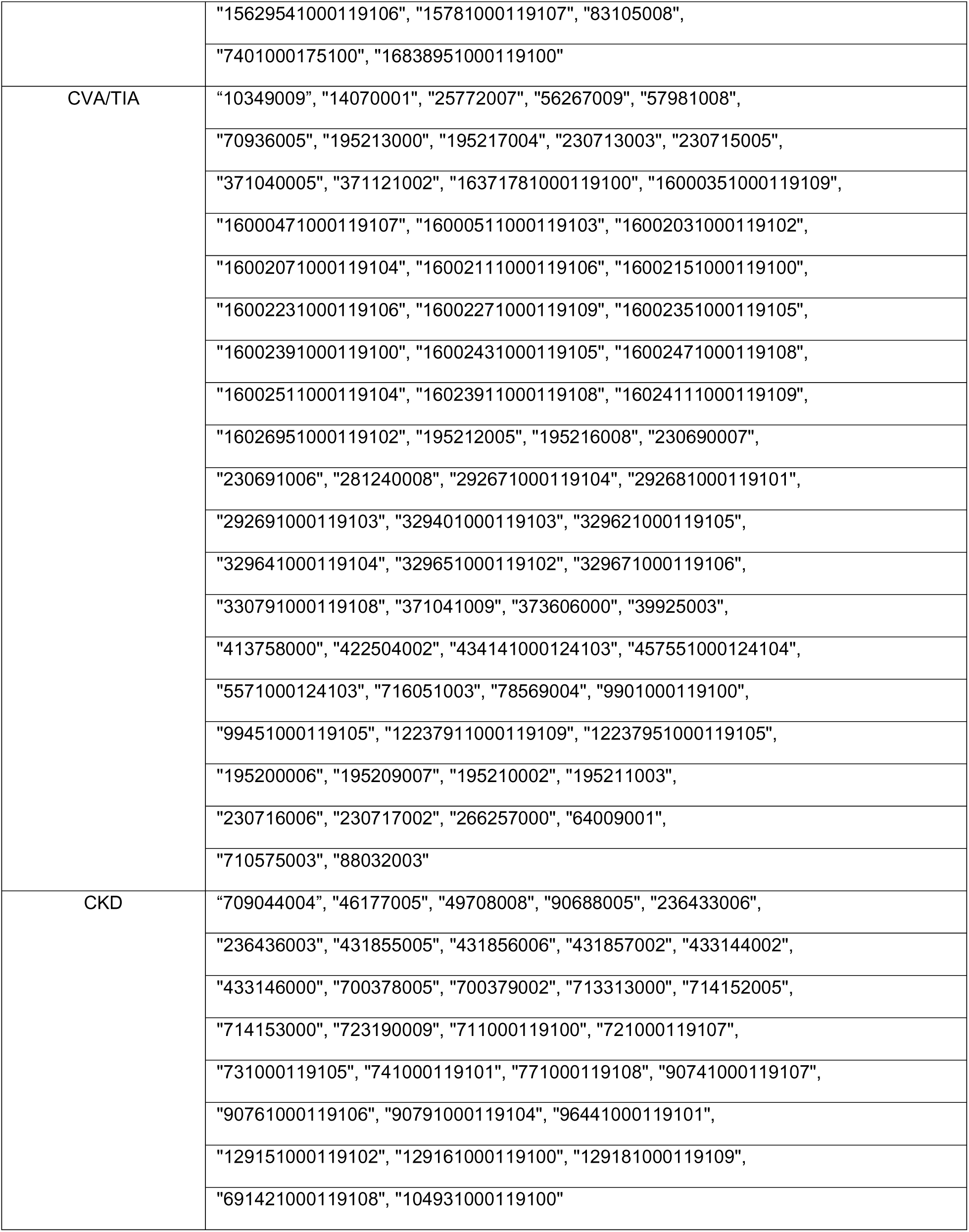
SNOMED CT codes used to identify each disease condition.

**Supplementary Table 2:** Mean age of demographic subgroups.

| Group | Subgroup | NHANES |  | AoU Survey |  | AoU EHR |  |
| --- | --- | --- | --- | --- | --- | --- | --- |
|  |  | Mean | SD | Mean | SD | Mean | SD |
| **Gender** | Female | 48.28 | 18.02 | 56.9 | 16.86 | 56.91 | 16.93 |
|  | Male | 46.65 | 17.67 | 62.6 | 17.04 | 60.63 | 16.78 |
| **Race/Ethnicity** | White | 49.79 | 18.12 | 61.41 | 16.82 | 62.13 | 17.04 |
|  | Black | 45.30 | 17.23 | 56.84 | 14.78 | 56.91 | 14.51 |
|  | Asian | 45.05 | 16.75 | 48.23 | 16.51 | 50.4 | 16.92 |
|  | Hispanic | 41.56 | 16.21 | 50.34 | 15.67 | 51.94 | 15.91 |
|  | Other | 46.04 | 16.92 | 49.59 | 16.73 | 50.58 | 16.95 |

**Supplementary Table 3.**
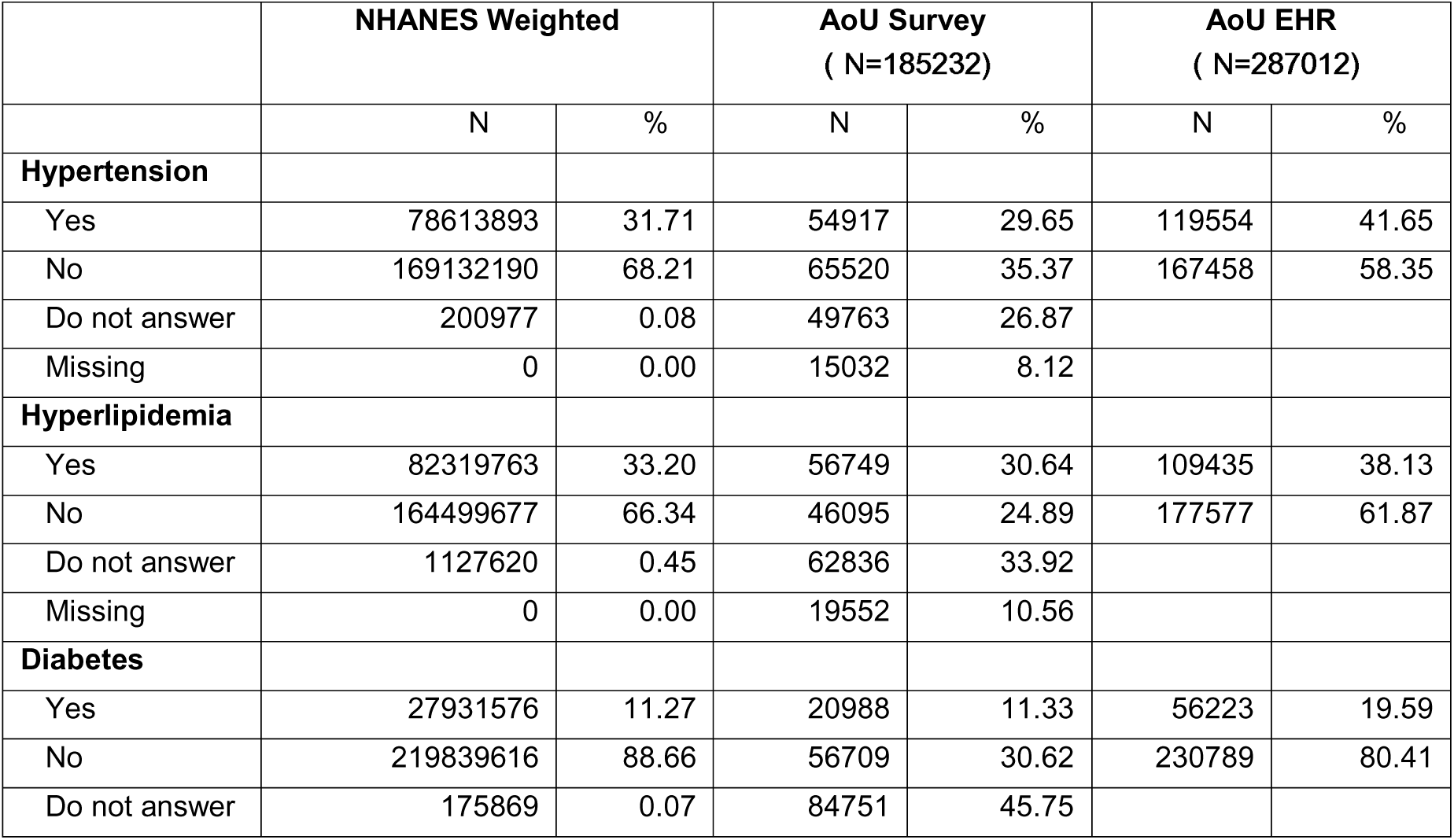
Prevalence of CV risk factors and established CVD.

**Supplementary Table 4.**
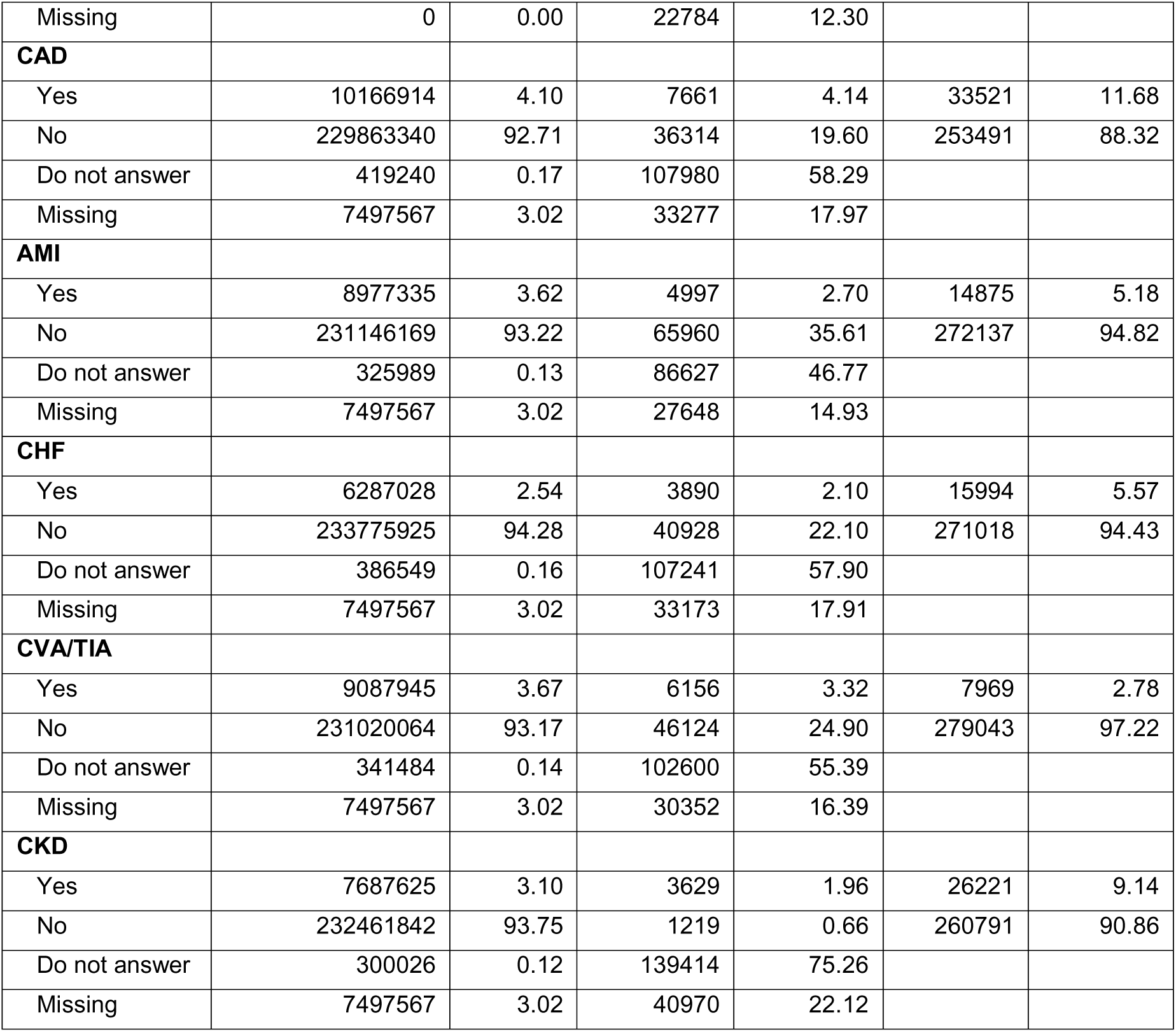
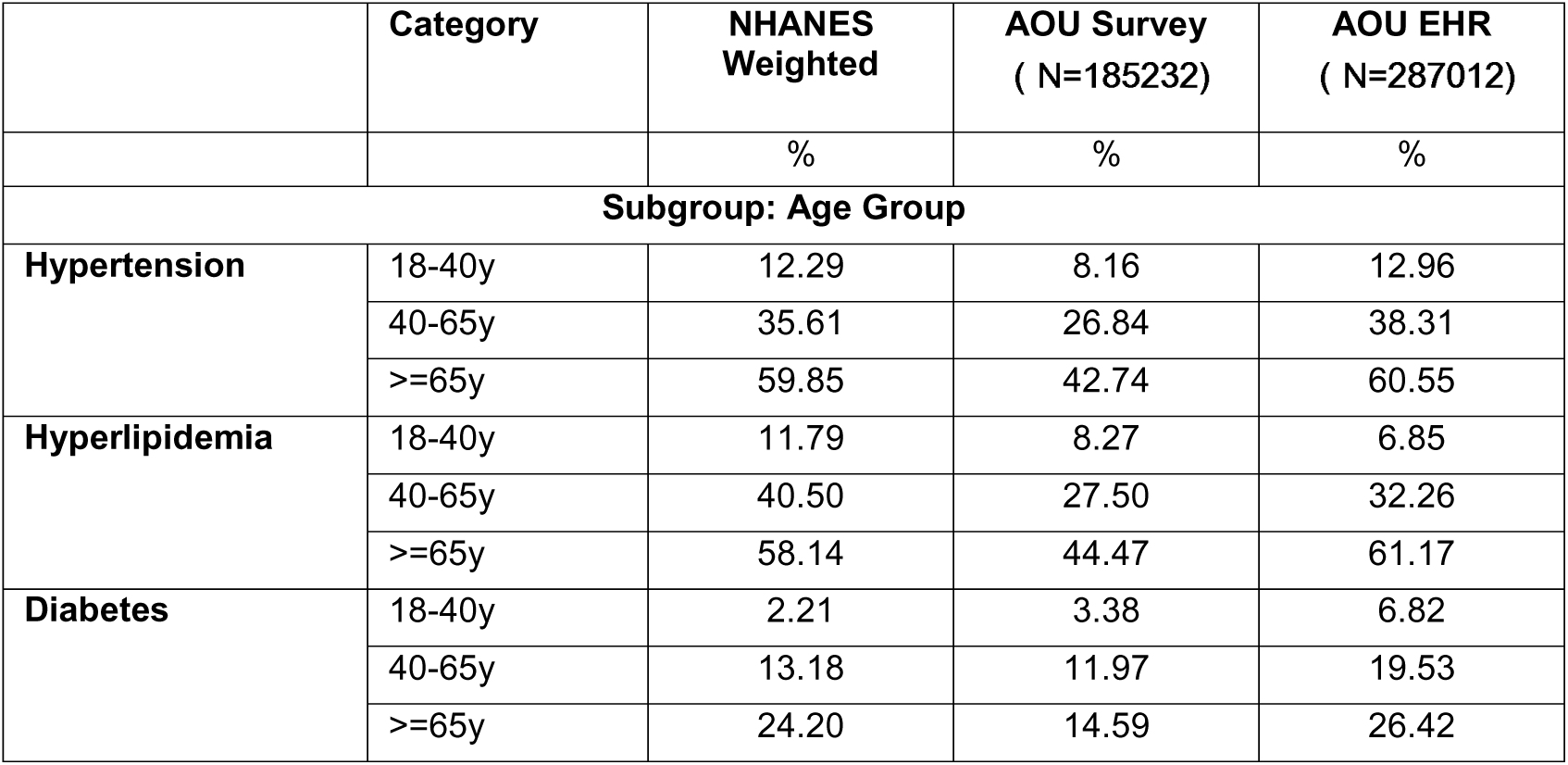
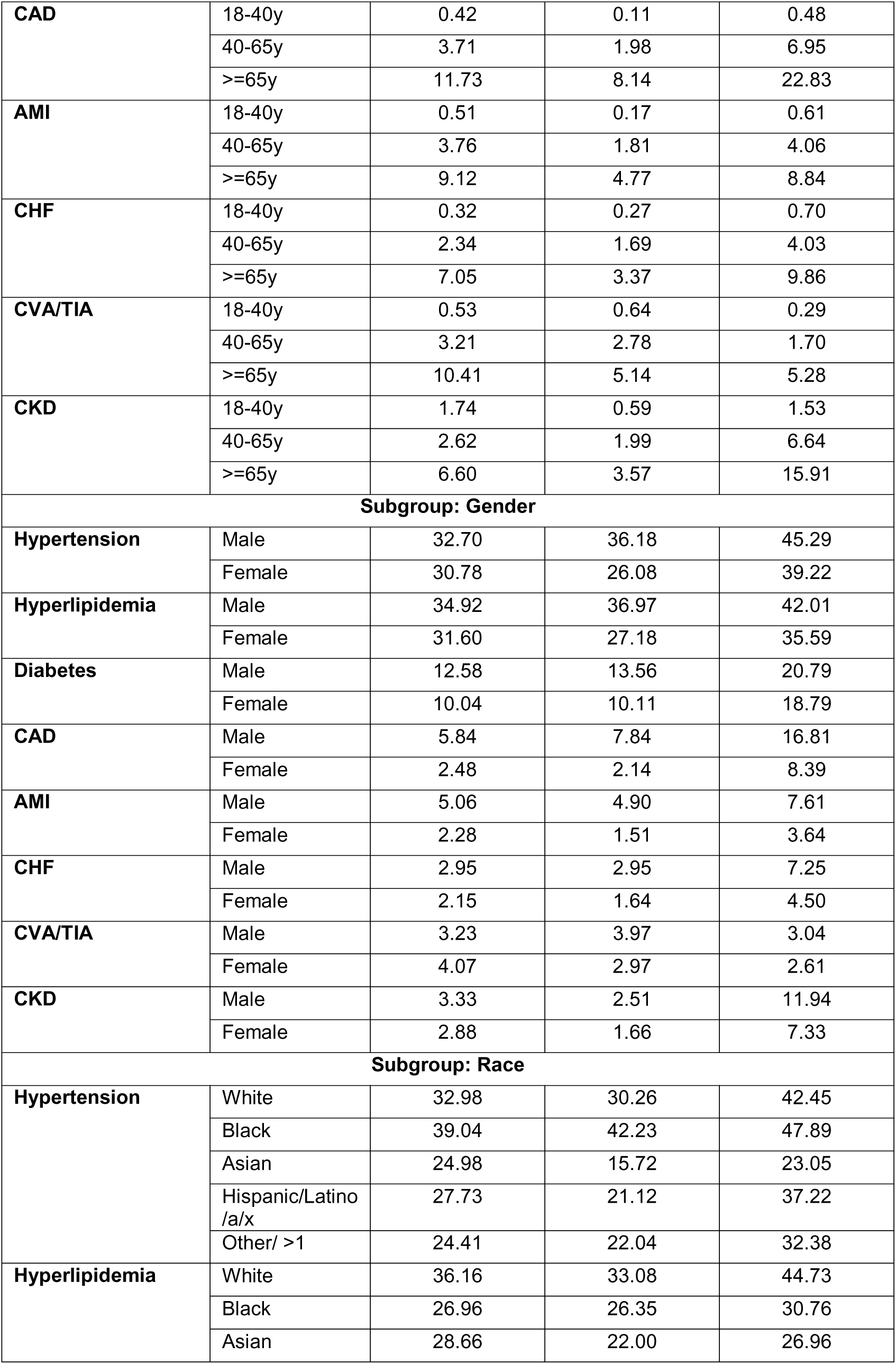
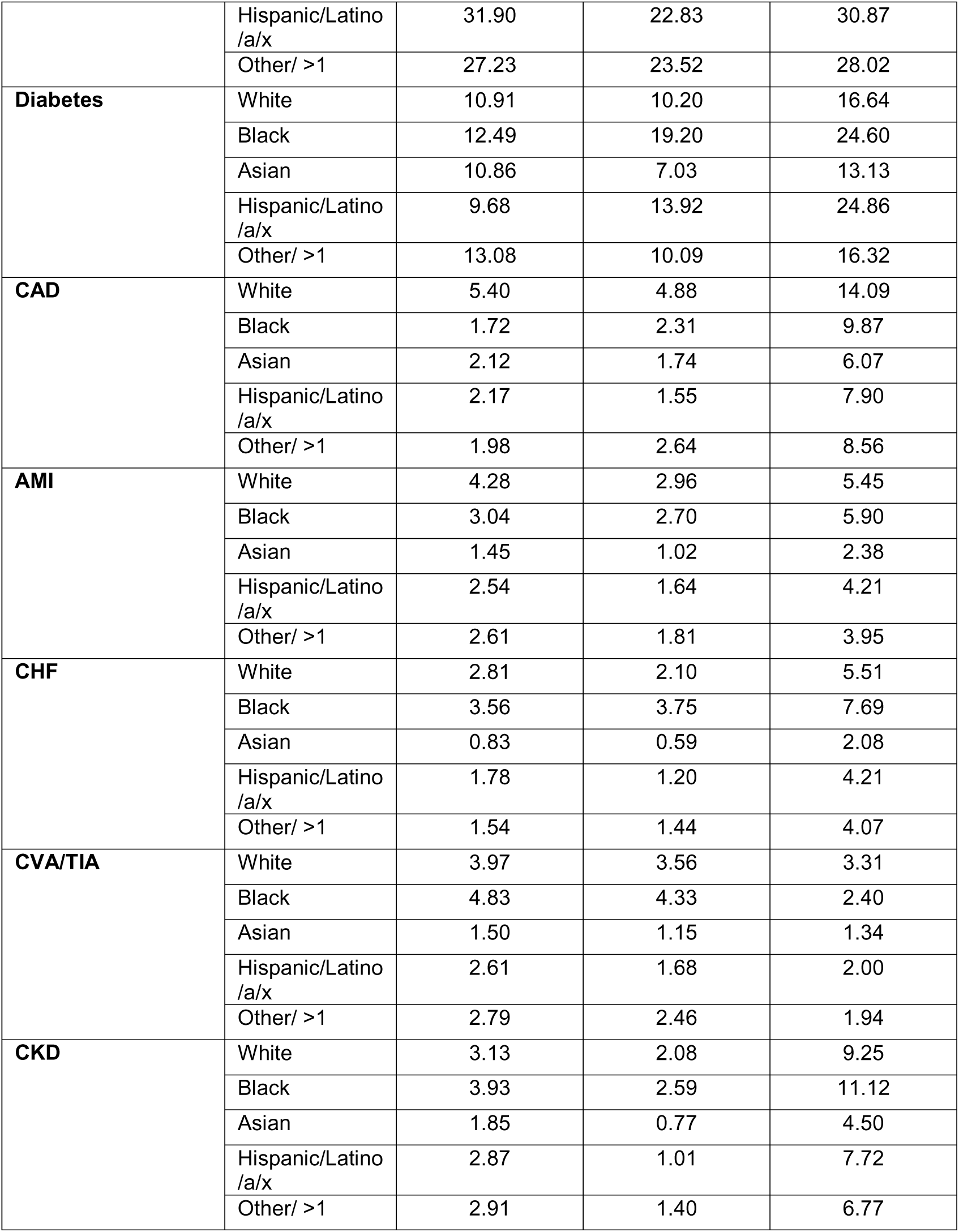
Prevalence of CV risk factors and established CVD by demographic subgroups.

